# Where should new parkrun events be located? Modelling the potential impact of 200 new events on geographical and socioeconomic inequalities in access and participation

**DOI:** 10.1101/19004143

**Authors:** PP Schneider, RA Smith, AM Bullas, T Bayley, SSJ Haake, A Brennan, E Goyder

## Abstract

**Background:** parkrun, an international movement which organises free weekly 5km running events, has been widely praised for encouraging inactive individuals to participate in physical activity. Recently, parkrun received funding to establish 200 new events across England, specifically targeted at deprived communities. This study aims to investigate the relationships between geographic access, deprivation, and participation in parkrun, and to inform the planned expansion by proposing future event locations.

**Methods:** We conducted an ecological spatial analysis, using data on 455 parkrun events, 2,842 public green spaces, and 32,844 English census areas. Poisson regression was applied to investigate the relationships between the distances to events, deprivation, and parkrun participation rates. Model estimates were incorporated into a location-allocation analysis, to identify locations for future events that maximise deprivation-weighted parkrun participation.

**Results:** The distance to the nearest event (in km) and the Index of Multiple Deprivation (score) were both independently negatively associated with local parkrun participation rates. Rate ratios were 0.921 (95%CI = 0.921-0.922) and 0.959 (0.959-0.959), respectively. The recommended 200 new event locations were estimated to increase weekly runs by 6.9% (from 82,824 to 88,506). Of the additional runs, 4.1% (n=231) were expected to come from the 10% most deprived communities.

**Conclusion:** Participation in parkrun is wide spread across England. We provide recommendations for new parkrun event location, in order to increase participation from deprived communities. However, the creation of new events alone is unlikely to be an effective strategy. Further research is needed to study how barriers to participation can be reduced.

**Online Map, data, and source code:** An interactive online map is available [here](https://iolmap.shinyapps.io/parkrun/), and the annotated R source code and all data that were used to generate the results of this study are provided on a repository.

## 1. Introduction

Insufficient physical activity is a leading cause of disease and disability world-wide.[1] For the UK, around one in six deaths is attributable to insufficient physical activity.[2] Increasing the physical activity levels of the population has the potential to improve quality of life, reduce mortality rates and alleviate the strain on health and social care services. However, interventions designed to increase population physical activity risk failing to reach those from the most deprived communities, potentially worsening health inequities.[3,4]

parkrun, an international movement which organises free weekly 5km running events, has been widely praised as being successful in encouraging participation in individuals who were previously inactive.[5,6] Participants report that accessibility and inclusivity are among the main factors that contributed to their involvement.[7] Sport England recently announced funding to support the creation of 200 new parkrun events across England within three years, with the aim of increasing participation of individuals from lower socio-economic groups.[8]

This study aims to provide recommendations for greenspace sites on which to establish new parkrun events. We start with an analysis of the relationship between deprivation, distance, and parkrun participation. We then use the observed relationship to propose a simple algorithm for identifying future event locations, in order to improve participation from deprived communities.

## 2. Methods

### 2.1. Data and Measures

This study is an ecological analysis of the geographic and socio-economic disparities in participation in parkrun events in England. The observational period covered 49 weeks in 2018 (1st January to 10th December). All analyses were conducted on the level of Lower Layer Super Output Area (LSOA). LSOAs are census areas with a population of approximately 1,700, which divide England into 32,844 geographic units.

We collected and combined location (longitude and latitude) and attribute data of LSOAs with information on parkrun events and public green spaces. An overview of the relevant measures and the data sources is provided in Table 1.

**Table 1:** Variables and data sources.

| Variable | Definition and source |
| --- | --- |
| **LSOAs (n=32,844)** |  |
| Location | Locations LSOAs were defined by their population-weighted centroids, retrieved from ONS.[10] |
| Population | Total 2017 population, retrieved from ONS.[11] |
| Index of Multiple deprivation (IMD) | The IMD is a measure of relative deprivation. It combines 37 indicators from seven domains (income, employment, education and skills, health and disability, crime, housing and services, and living environment) into a single score, ranging from 0 (least deprived) to 100 (most deprived). Data taken from official statistics.[12,13] |
| Distance (in km) | The linear (geodesic) distance from a LSOA to the nearest parkrun event in km. To derive this figure, all distances between LSOAs and parkrun events (32,844 x 455 matrix) were computed. Subsequently, the shortest distance was selected for each LSOA. |
| Participation/Runs (runs per LSOA) | Participation for a given LSOA was measured as the number of times anyone living in the LSOA attended, i.e. did a run in an English parkrun event within the observational period of 49 weeks. Data provided by parkrun UK.[14] |
| Participation rate (runs per 1,000 per week) | To standardise participation across LSOAs, we computed the average number of runs per 1,000 population per week. |
| **parkrun events (n=455)** |  |
| Location | The locations of parkrun events were collected from the official parkrun website.[15] |
| **Green spaces (n=2,842)** |  |
| Location | Coordinates of public parks and gardens were retrieved from the openly available OS Open Greenspace dataset.[9] |
| Area | Area of the green space in km<sup>2</sup>. |

In this study, we included all 455 English parkrun events which were in operation during the observational period. Events held in prisons (n=7) were excluded. Data on 143,822 public green spaces were retrieved from an open dataset of Ordnance Survey.[9] In the absence of more detailed information (e.g. terrain), we considered all public parks, gardens, and playing fields in England with an area of 0.1 km^2^ (e.g. 316m x 316m) or more potentially suitable for hosting parkrun events (n=2,842).

### 2.2. Analysis

Mean, standard deviation, median, interquartile range, and range were used as descriptive statistics. The associations between IMD, distance to the nearest parkrun event, and parkrun participation on the LSOA level were investigated using generalised linear models with a Poisson distribution and a log link. The dependent variable was the total number of runs between 1st January and 10th December 2018. To model the outcome as a rate (runs per 1,000 population per week), the log of the person weeks was used as an offset variable.

We first fitted bivariate models to study the effect of IMD and distance on participation separately. The correlation between both predictors was assessed using Pearsons correlation coefficient. Subsequently, we used a multivariable model to investigate the independent effects of IMD and distance on participation. The Akaike (AIC) and the Bayesian Information Criterion (BIC), as well as the McFaddens pseudo R^2^ were used to assess model fit.

### 2.3. Identifying optimal locations for new parkrun events

#### 2.3.1. Rationale and objective

We conducted a location-allocation analysis to solve the following problem: parkrun UK received funding to start 200 additional parkrun events. There are 2,842 public green spaces in England in which new events could be set up. Which 200 locations should be selected?

parkruns stated aim of increasing participation from lower socio-economic groups[8] was operationalised in the following way: *find the set of 200 green spaces, which maximise the number of additional runs at parkrun events, weighted by LSOAs squared IMD scores*. By using the squared IMD scores as weights, the generation of new runs from more deprived areas is prioritised over new runs from less deprived areas. We also explored the following alternative specifications: 1. minimise deprivation-weighted distances; 2. maximise the total (i.e. unweighted) number of runs; 3. Minimise (unweighted) distances.

#### 2.3.2. Location-allocation analysis

We developed a flexible algorithm to select the optimal new parkrun event locations from the set of candidate green spaces. The algorithm consists of two steps: firstly, for each green space, we evaluate how many new runs (weighted by squared LSOA IMD score) would be generated by setting up a new parkrun event at the respective location. Secondly, the green space with the greatest effect is selected. This procedure is repeated 200 times.

More formally, we define that for any candidate green space location j, the objective function *f* (*j* | *E*) provides the sum of parkrun runs *r*_*i*_ over all LSOA *i*, weighted by the squared IMD score 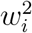, given the set of established parkrun event locations *E* = {*e*_1_, *e*_2_,“, *e*_455_}:

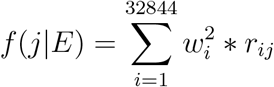

In the absence of causal estimates, we use the Poisson regression model specified above to predict the expected number of runs *r*_*ij*_ for LSOA *i* based on its IMD score *w*_*i*_, its (linear) distance to the nearest parkrun event *d*_*ij*_, and its population *p*_*i*_. The functional form is given below.

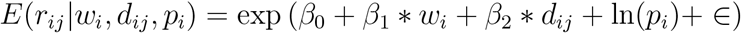

Filling-in the parameter coefficients (see table 3), we derive the following formula:

**Table 2:** Descriptive statistics for LSOA and parkrun event attributes.

| **LSOA (n =n 32,844)** |  |  |  |
| --- | --- | --- | --- |
|  | Mean (SD) | Median (Q<sub>1</sub>-Q<sub>3</sub>) | Range |
| Population | 1,693.44 (405.35) | 1,612 (1452-1834) | 362-13404 |
| IMD score | 21.67 (15.59) | 17.40 (9.65-30.07) | 0.48-92.60 |
| Distance to the nearest parkrun event location | 4.74 (4.26) | 3.46 (2.02-5.95) | 0.04-76.44 |
| Total runs per LSOA to English parkrun events* | 123.57 (128.89) | 86 (33-172) | 0-1659 |
| Runs per 1,000 LSOA population per week | 1.52 (1.55) | 1.07 (0.41-2.12) | 0.00-16.48 |
| **Parkrun Events (n = 455)** |  |  |  |
|  | Mean (SD) | Median (Q<sub>1</sub>-Q<sub>3</sub>) | Range |
| Served LSOAs<sup>†</sup> | 72.18 (44.07) | 63(40-89) | 6-350 |
| Served population (1,000s)<sup>†</sup> | 122,240.5 (76,380.64) | 106,228 (69,318-154,778.5) | 7,855-628,010 |
| Mean participants per event per week | 188.17 (96.15) | 170.50 (117.85 -245.65) | 32.80-682.80 |

\* Runs to English parkrun events between 1st Jan – 10<sup>th</sup> Dec 2018; † Number of LSOAs or population for which a given parkrun event is the nearest.

**Table 3:** Poisson models for the bivariate and multivariable effects of distance and deprivation on Parkrun participation rates per LSOA. Total population and time were used as offset variables.

|  | Distance model<br>Rate ratio (95%CI) | IMD model<br>Rate ratio (95%CI) | Multivariable model<br>Rate ratio (95%CI) |
| --- | --- | --- | --- |
| Intercept | 0.002 (0.002; 0.002) | 0.003 (0.003; 0.003) | 0.005 (0.004; 0.005) |
| Distance<sup>†</sup> | 0.943 (0.943; 0.944) |  | 0.921 (0.921; 0.922) |
| IMD |  | 0.962 (0.962; 0.962) | 0.959 (0.959; 0.959) |
| AIC | 3751473 | 2973296 | 2655185 |
| BIC | 3751490 | 2973312 | 2655210 |
| McFadden R<sup>2</sup> | 0.05 | 0.25 | 0.34 |

AIC = Akaike Information Criterion; BIC = Bayesian Information Criterion; IMD = Index of Multiple Deprivation; †Distance = Linear geodesic distance to the nearest parkrun event.

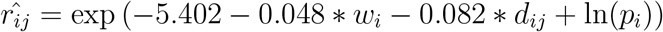

Note that *j* can have an effect on *r*_*ij*_ through *d*_*ij*_ : setting up a new event at location *j* will reduce the distance to the nearest event for some LSOA *i*. This means, we evaluate the distances from LSOA *i*s location *l*_*i*_ to all established parkrun event locations {*e*_1_, *e*_2_, “, *e*_455_} ∈ *E*, denoted 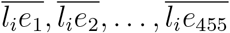, and to the candidate green space location *j*, denoted 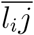, and then take the minimum value, i.e. 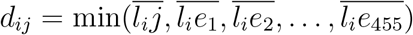.

The expected change in the objective function is computed for all candidate locations j in the set of the available green spaces *C* = {*c*_1_, *c*_2_, “, *c*_2842_}, and the location with the maximum value is selected. The selection function is expressed in the following formula:

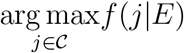

In order to identify the optimal set of 200 new locations *O* = {*o*_1_, *o*_2_, “, *o*_200_}, the selection procedure is repeated 200 times. At each step *k*, the best candidate green space location *o*_*k*_ ∈ *C* is selected, added to the set of established parkrun events *E*, and removed from the set of available green spaces *C*. Thereby, the *k*th new parkrun event location is taken into account when selecting the *k*th+1 location. The full algorithm can be described in pseudo code in the following way:

### 2.4. Interactive map, source code and data

We mapped all relevant data points, including LSOAs, green spaces, parkrun events, and recommended new event locations, and created an interactive map, which can be accessed online [16]. An annotated version of the R source code and all data that were used to generate the results of this study are provided on a repository under a creative commons license [17].

### 2.5. Ethical approval

Ethical approval was obtained from the Sheffield Hallam University Ethics Committee (ER10776545). We did not collect any personal information, but only used aggregate secondary data. The parkrun Research Board approved this research project, and three of its members (AMB; EG, SSJH) were actively involved in it.

## 3. Results

### 3.1. Descriptive statistics

As of 10th December 2018, approximately 7%, 69%, and 90% of the English population lived within 1, 5, and 10km of a parkrun event. Only 192,208 people (0.35% of the English population) lived more than 25km from their nearest event. Over the study period of 49 weeks, an average of 82,824 participants attended parkrun events each week. This is equivalent to a national average weekly participation rate of 1.49 per 1,000 population (this corresponds to an unweighted average of 1.52 per LSOA). About 3% of these runs were done by runners living in the 10% most deprived LSOAs, while 18% came from the 10% least deprived LSOAs. Further descriptive statistics are provided in Table 2.

### 3.2. Relationship between deprivation, distance and participation in parkrun events

Table 3 shows the results of the bi- and multivariable Poisson regression models. We found that the distance to the nearest event and the IMD score were both negatively associated with a LSOAs parkrun participation rate. LSOAs with a further distance to the nearest parkrun event, and those with a high IMD score (i.e. more deprived) had lower parkrun participation rates. The reported adjusted rate ratios denote the expected relative change in the average number of runs from a one unit increase in the predictor variable. After controlling for IMD, an increase in the distance to the nearest parkrun event by 1km was associated with a 7.9% lower parkrun participation rate (adjusted rate ratio = 0.921). The adjusted rate ratio for IMD was 0.959, respectively. Due to the high number of observations (n=32,844), confidence intervals were very narrow, with a width of less than 0.01 for all coefficients. The collinearity between distance and IMD was low (Pearson correlation coefficient *r* = −0.14), and their effect sizes were similar across the bivariate and the multivariable Poisson regression models, which suggests that both predictors explained different parts of the variance. Moreover, models goodness-of-fit values suggested that IMD scores were markedly better than distance to the nearest event in predicting participation (McFadden Pseudo R^2^= 0.05 vs. 0.25).

Figure 1 illustrates the results of the multivariable regression model. Each point shows the observed distance to the nearest event and the number of runs per 1,000 population per week for a single LSOA (n=32,844). We plotted the estimated relationship between distance and participation for three IMD levels: the 10th percentile (least deprived; IMD = 5.67), 50th percentile (median; 17.40) and 90th percentile (most deprived; 44.57).

**Figure 1:**
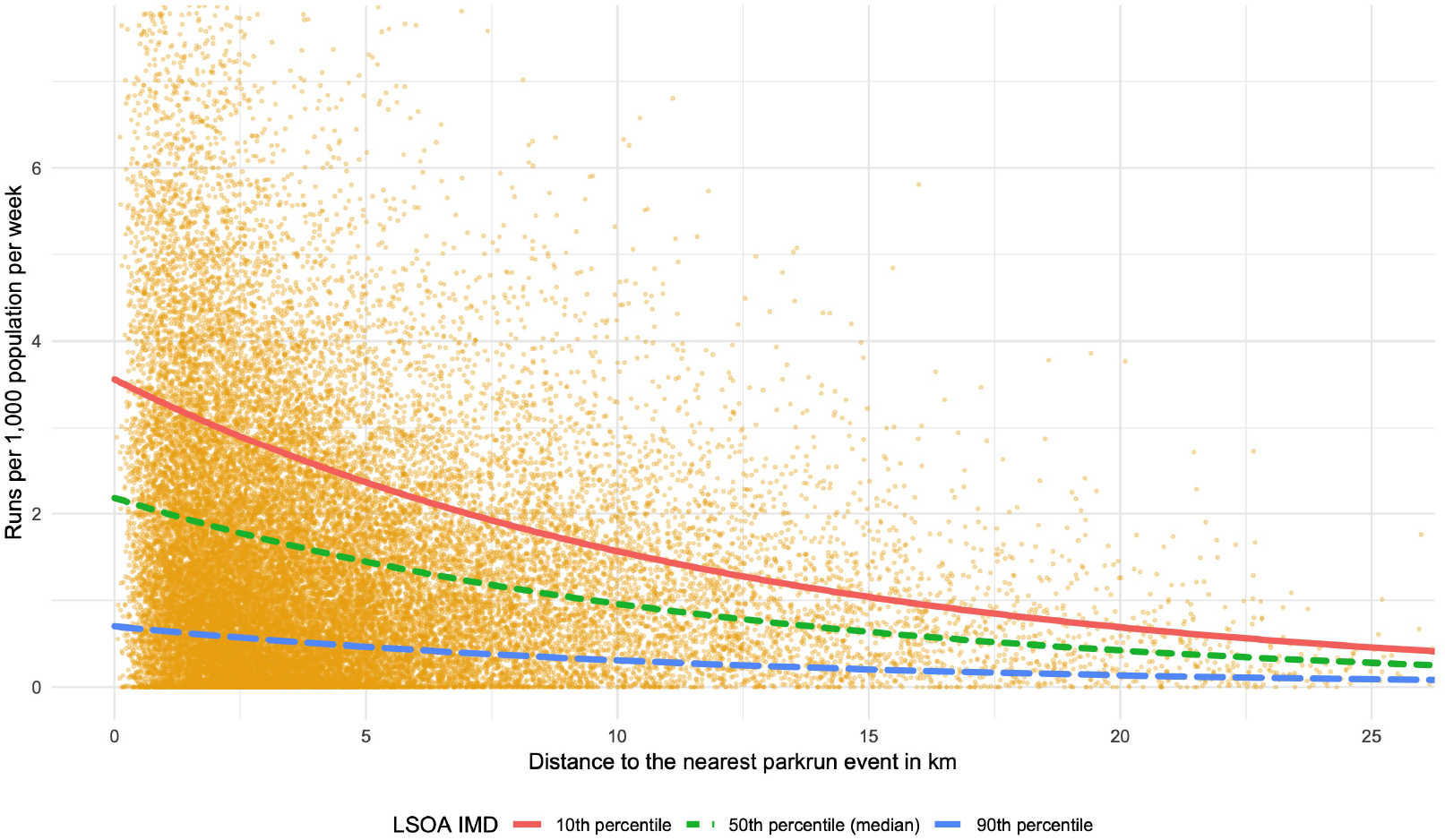
Observed mean weekly parkrun participation rate by distance to nearest parkrun event for each LSOA (points), and model estimates for the 10th, 50th and 90th IMD percentiles (lines).

As can be seen from the graphs, participation rates for the least deprived LSOAs are markedly higher, compared to the most deprived areas. At all levels of deprivation, participation falls as distance to nearest parkrun event increases. However, the most deprived LSOAs are much less responsive to changes, and their participation rates remain relatively low for any given distance.

### 3.3. Optimal locations for new parkrun events

Figure 2 shows current English parkrun events (black circles) alongside recommendations for 200 additional event locations (red triangles), which maximise the total parkrun participation, weighted by the squared IMD score. The numbers correspond to the rank, where 1 is the location which would improve the weighted participation the most. The names and exact locations of the selected green spaces are provided in the appendix.

**Figure 2:**
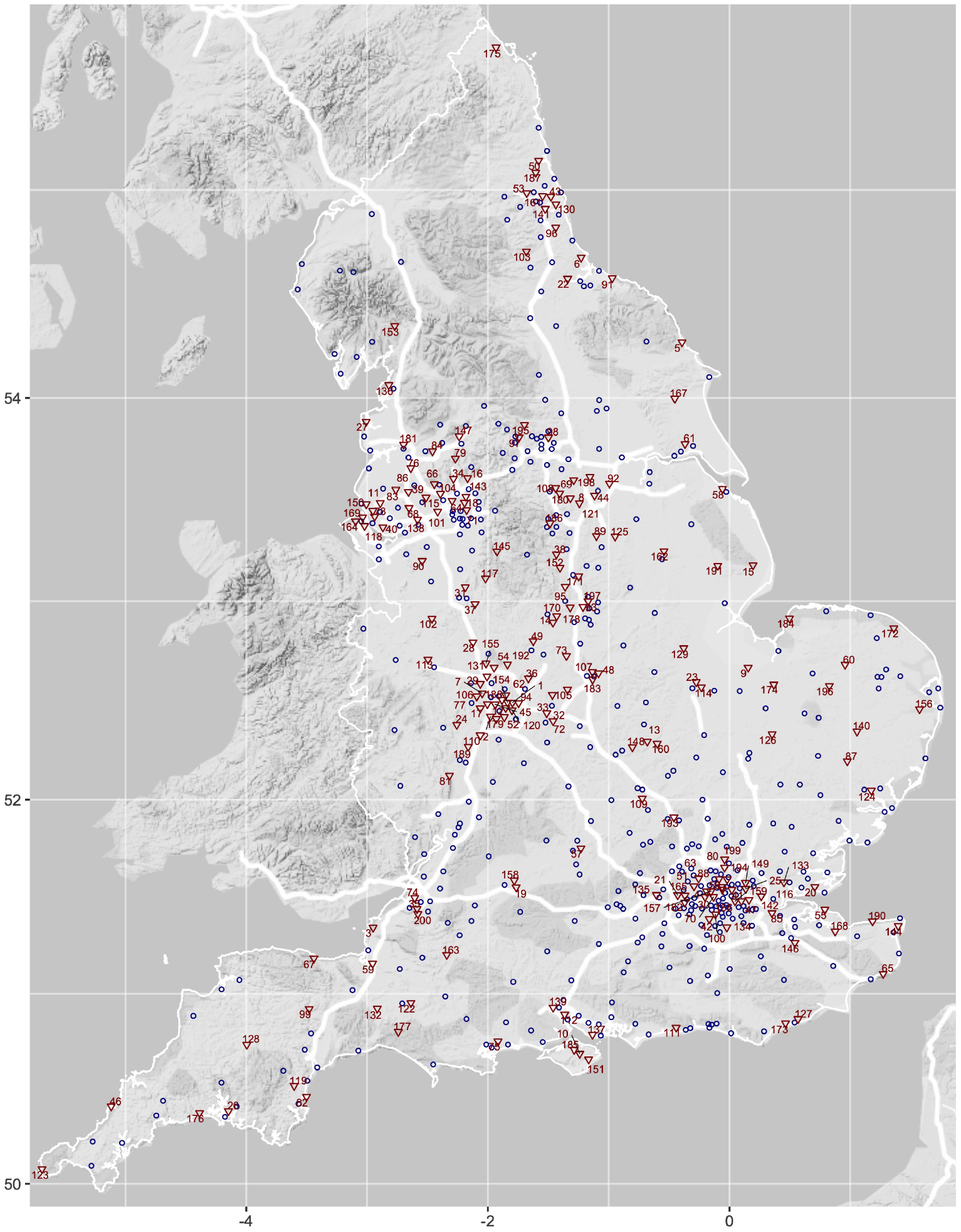
Map of England showing current parkrun events (blue circles) and recommended new event locations (red triangles) ranked in descending order of estimated effect on IMD weighted participation. A legend, listing all 200 recommended new parkrun event locations, is provided in the appendix.

We estimated that the 200 new events would reduce the average distance to the nearest parkrun event by 1.15km (SD = 2.71, range = 0.00-47.55).

The (potential) geographic access to parkrun would be improved for about 18.3 million people, i.e. 33% of the population. Based on the multivariable regression model (see table 3), we estimate that the 200 new events would result in an additional 5,682 runs per week, an increase of about 7%. Of these new runs, we estimated that 231 (4%) originate from the 10% most deprived LSOAs.

In addition to the results reported here, we also explored alternative objective specifications (1. maximising total participation, 2. minimising total distances, 3. minimising squared-IMD-weighted distances). For the location recommendations which maximise the (unweighted) total participation, we estimated that a 33% higher number of weekly runs could be achieved (n = 7,530). However, fewer runs originated from the 10% most deprived LSOAs (n=110), while 14 times more runs originated from the 10% least deprived LSOAs (n=1,493). Detailed results for alternative objective specifications are provided in the appendix. In addition, an interactive map, displaying the recommendations alongside existing parkrun events is provided online [16].

## 4. Discussion

### 4.1. Main findings

Our study supports parkruns planned expansion by providing recommendations for the optimal 200 new parkrun event locations. Based on our multivariate regression model, we estimate that the new events would reduce the average distance to the nearest parkrun event by 1.15km, and generate an additional 295,444 runs per year, which would be an increase of 7%, compared to the average participation in 2018.

However, our policy recommendations are expected to have only a modest effect on the participation from the most deprived LSOAs. We found that participation rates from these communities are low, even when geographical access is very good. Improving geographic access, by creating new events, is therefore unlikely to substantially increase the number of participants from lower socio-economic groups. In fact, setting up new parkrun events is likely to worsen inequalities in participation. Therefore, complementary measures should be considered.

Notwithstanding the above, it should be noted that in 2018, participation in parkrun events was widespread across England: on average, 82,824 people, that is 0.15% of the population participated in parkrun events each week, with participants coming from 95% of all LSOAs. Geographic access can be assumed to be an enabling factor for their success: about 69% of the English population live within 5km of a parkrun event. Interestingly, we observed a weak negative correlation between a LSOAs IMD and the distance to the nearest event (r = −0.14), suggesting that parkrun events tend to be located closer to more deprived communities.

Optimal locations for new parkrun events are contingent on how the objective function is specified. The recommendations reported here aim to maximise the total number of parkrun runs, weighted by the squared IMD score. This reflects a strong preference towards increased participation from deprived communities - a run from a LSOA in the 90th IMD percentile (most deprived) had 62 times the weight of a run from the 10th percentile (least deprived). However, alternative specifications may also be legitimate: without IMD-weights, for example, it would be possible to generate a markedly higher number of total runs. Moreover, one could argue that parkrun should not seek to maximise actual participation, but rather focus on giving people the opportunity to participate [18], by minimising distance to nearest event corresponding location recommendations are provided in the appendix. The decision where new events should be located involve normative judgements, which will depend on the values and preferences of the decision maker. This study is not meant to provide definite location recommendations, but to showcase a potentially useful decision aid. To enable parkrun and other stakeholders to adjust the objective function and repeat the analysis, we provide the source code and data that were used for this study.

### 4.2. Limitations

In our analysis, we had to make several simplifying assumptions that should be considered when interpreting our findings. Firstly, the use of linear (geodesic) distances is likely to underestimate true travel distance. Secondly, included candidate park locations that may not necessarily be suitable for parkrun (e.g. due to a lack of paths). Likewise, it is possible that some suitable sites, for example green spaces not categorised as public parks, were not included in the study.[17]

Furthermore, we conducted an ecological analysis on the level of the LSOA. Since there might be substantial variation within communities, further studies are required to confirm whether the estimated relationships hold at the individual level.[19]

Finally, it is important to note that, although we used the multivariable regression analysis (see Table 3) to predict the effect of setting up new parkrun events on LSOA participation rates, the models were not specified to investigate causal relationships [20,21]. There might be other important factors that explain participation [7] - the generally increasing popularity of parkrun across England, for example - that were not considered. However, building a causal model to reliably predict future parkrun participation would require more detailed and longitudinal data, as well as more extensive modelling.

### 4.3. Implications for policy

This is the first study to investigate the socio-economic and geographic disparities in participation in parkrun events in England. We aimed to support parkruns planned expansion by identifying green space sites for new events which maximise participation, while incorporating concerns about socio-economic equity. However, our findings call into question whether the creation of new events is the most effective strategy to increase participation from lower socio-economic groups. parkrun might want to consider how they could complement the current efforts to improve geographic access to parkrun events, in order to encourage people from deprived communities to participate.

Further research is required to identify and reduce the barriers for participation in parkrun events, especially for people living in deprived areas. For this purpose, events that are particularly successful in recruiting people from deprived areas could serve as learning opportunities: deep-dive qualitative research at these events may improve understanding of how parkrun events can better reach the targeted population groups.

### 4.4. Conclusion

We provide suggestions for optimal locations of new parkrun events to maximise participation from deprived communities. The creation of new events alone, however, might be an ineffective strategy. In fact, since less deprived communities generally have higher participation rates, the creation of new events might even worsen existing inequities. More research is needed to identify effective strategies to increase participation of individuals from low socio-economic groups.

## Data Availability

All data and the R source code that were used to generate the reported results are openly provided on a github repository under a CC BY license.

https://github.com/bitowaqr/iolmap_analysis

https://github.com/bitowaqr/iol_map

http://iol-map.shef.ac.uk/

## Acknowledgements

The authors would like to thank the parkrun research board for their support. We are also grateful to parkrunUK for the collection, management and provision of the parkrun data, and for meeting with us to discuss preliminary work. parkrunUK did not have an active role in the study, and all opinions and interpretations of the results are those of the authors, and do not necessarily reflect the views of parkrunUK or the parkrun research board. This work was supported by the Wellcome Trust through a PhD studentship.

